# SARS-CoV-2-specific Humoral and Cell-mediated Immune Responses after Immunization with Inactivated COVID-19 Vaccine in Kidney Transplant Recipients (CVIM 1 Study)

**DOI:** 10.1101/2021.08.02.21261095

**Authors:** Jackrapong Bruminhent, Chavachol Sethaudom, Pongsathon Chaumdee, Sarinya Boongird, Sasisopin Kiertiburanakul, Kumthorn Malathum, Arkom Nongnuch, Angsana Phuphuakrat, Sopon Jirasiritham, Chitimaporn Janphram, Sansanee Thotsiri, Supparat Upama, Montira Assanatham, Ramathibodi Transplant Infectious Diseases (RTID) Study Group

**Author notes:** These authors contributed equally to this work (Jackrapong Bruminhent, Chavachol Sethaudom). **Corresponding author:** Montira Assanatham, MD, Division of Nephrology and the Excellence Center for Organ Transplantation, Department of Medicine, Faculty of Medicine Ramathibodi Hospital, Mahidol University, 270 Rama VI Road, Ratchathewi, Bangkok, 10400, Thailand.

## Abstract

Immunogenicity following inactivated SARS-CoV-2 vaccination among solid organ transplant recipients has not been assessed. Seventy-five patients (37 kidney transplant [KT] recipients and 38 non-transplant controls) received two doses, at 4-week intervals, of an inactivated whole-virus SARS-CoV-2 vaccine. SARS-CoV-2-specific humoral (HMI) and cell-mediated immunity (CMI) were measured before, 4 weeks post-first dose, and 2 weeks post-second dose. The median age of KT recipients was 50 years (IQR, 42–54) and 89% were receiving calcineurin inhibitors/mycophenolate/corticosteroid regimens. The median time since transplant was 4.5 years (IQR, 2–9.5). Among 35 KT patients, anti-RBD IgG titer after vaccination was not significantly different to baseline, but was significantly lower than in controls (7.8 [95%CI 0.2–15.5] vs 2,691 [95%CI 1,581–3,802], p<0.001) as well as the percentage of surrogate virus neutralizing antibody inhibition (2 [95% CI −1-6] vs 71 [95%CI 61–81], p<0.001). However, the mean of SARS-CoV-2 mixed peptides-specific T-cell responses measured by enzyme-linked immunospot assays was significantly increased compared with baseline (66 [95%CI 36–99] vs. 34 [95%CI 19–50] T-cells/10^6^ PBMCs, p=0.02) and comparable to that in controls. Our findings revealed weak HMI and marginal CMI responses in fully vaccinated KT recipients receiving inactivated SARS-CoV-2 vaccine. (Thai Clinical Trials Registry, TCTR20210226002).

## 1. INTRODUCTION

Severe acute respiratory syndrome coronavirus 2 (SARS-CoV-2) is a recently-emerged pathogen causing coronavirus disease 2019 (COVID-19), which spread worldwide. The clinical manifestations vary from asymptomatic to mild upper or severe lower respiratory tract disease.^1^ Solid-organ transplant (SOT) recipients are among those who are potentially compromised for this particular infection, resulting in significant morbidity and substantial mortality in this demographic.^2–5^ Vaccination against SARS-CoV-2 is recommended to ameliorate this potentially serious infection and its unfavorable consequences. Various COVID-19 vaccines have been developed across a range of platforms and have been deployed among immunocompetent individuals. However, immunogenicity and safety data following COVID-19 vaccination among SOT recipients receiving immunosuppressants remain limited.

A messenger RNA (mRNA)-based COVID-19 vaccine has been shown to produce immune responses and adequate efficacy to prevent natural infection in immunocompetent recipients.^6,7^ However, recent studies focusing on immunogenicity following a two-dose, 4-week interval mRNA-based COVID-19 vaccination strategy revealed suboptimal immune responses among immunocompromised patients. Only 17% and 54% of participants generated robust immune responses after single and double doses, respectively, of the mRNA-based COVID-19 vaccine.^8–10^ An inactivated SARS-CoV-2 vaccine has been shown to be primarily adequate to prevent death (86% efficacy), with reportedly lower effect against disease acquisition (65.9%).^11^ However, a study focusing on immunogenicity and safety following vaccination with an inactivated whole-virus SARS-CoV-2 vaccine among SOT recipients has not been assessed.^12^ Furthermore, safety concerns for these immunocompromised patients have not been investigated.

Herein, we decided to conduct an immunogenicity study among kidney transplant (KT) recipients following a full course of inactivated SARS-CoV-2 vaccine. Both SARS-CoV-2 specific humoral (HMI) and cell-mediated immune (CMI) responses were investigated along with the safety profile.

## 2. MATERIALS AND METHODS

### 2.1 Study design

Between April 2021 and July 2021, we performed a prospective cohort study of adult KT recipients who received a two-dose, 4-week interval vaccination with an inactivated whole-virus SARS-CoV-2 vaccine, CoronaVac^®^ (Sinovac Biotech Ltd., China), which contains 3 μg of inactivated whole-virus SARS-CoV-2 in 0.5 mL, given intramuscularly into the deltoid muscle. We included KT recipients from three hospitals; Ramathibodi Hospital, Praram 9 Hospital, and Samitivej Sukhumvit Hospital, all located in Bangkok, Thailand.

HMI and CMI were measured before, 4 weeks after the first dose, and 2 weeks after the second dose, using a SARS-CoV-2 immunoglobulin G (IgG) assay that tests for antibodies against the receptor-binding domain (RBD) of the SARS-CoV-2 spike protein, SARS-CoV-2 surrogate virus neutralization test (sVNT), and an enzyme-linked immunospot (ELISpot) assay for interferon-γ (IFN-γ), respectively (**Supplementary Figure 1**).

Participants were eligible if they were KT recipients aged 18 to 59 years old, at least 1 month post-transplant, and stable in their allograft function and immunosuppressive regimen. In addition, patients with suspected respiratory tract infection in the preceding 3 days, concurrent active infection, recent diagnosis of allograft rejection requiring intense immunosuppressants (methylprednisolone pulse therapy with 500 mg IV daily for 3 days, antithymocyte globulin therapy within 3 months, rituximab therapy within 6 months, or prednisolone more than 15 mg/day), receiving other vaccination within 4 weeks, previous history of COVID-19, or prior administration of COVID-19 vaccine were excluded. All included patients were screened for active respiratory tract infection, recent COVID-19 exposure, and comorbidities by history. Nasopharyngeal and oropharyngeal swabs for SARS-CoV-2 reverse transcription polymerase chain reaction (RT-PCR) were not performed before vaccination. Thirty-eight non-transplant adult patients who did not receive immunosuppressants were included and referenced as a control. They also received the same type and interval of COVID-19 vaccination and were assessed for immunity as described above.

### 2.2 SARS-CoV-2 humoral immune responses

SARS-CoV-2 anti-RBD IgG antibodies were measured using the Abbott SARS-CoV-2 IgG II Quantification assay (Abbott Diagnostics, U.S.A). Plasma samples were run on the Abbott Alinity instrument following the manufacturer’s instructions. The assay is a chemiluminescent microparticle immunoassay for the quantitive detection of IgG in human serum against the RBD of the SARS-CoV-2 spike protein. The quantitative results of anti-RBD IgG were reported in arbitrary units (AU)/mL.

The function of the anti-SARS-CoV-2 spike protein S1 RBD antibody was determined by using a SARS-CoV-2 NeutraLISA surrogate neutralization test assay (Euroimmun, Germany). The neutralizing antibodies in plasma were inhibit binding between RBD and angiotensin-converting enzyme 2 (ACE2) receptor. The percentage of neutralizing antibody inhibition was reported.

### 2.3 SARS-CoV-2-specific cell-mediated immune responses

According to the manufacturer’s protocol, heparinized whole blood samples from participants were collected, and peripheral blood mononuclear cells (PBMCs) were isolated using the EasySep™ Direct Human PBMC Isolation Kit (Stemcell Technologies, Vancouver, Canada). Isolated cells were counted, and the cell suspension was normalized at a final concentration of 2.5×10^6^ cells/mL in AIM V media (Gibco, Waltham, MA) followed by manually plating of cells into strip plates (2.5×10^6^ PBMCs/well) for stimulation with peptide pool or cell stimulation cocktail.

ELISpot assays assessed IFN-γ production by activated PBMCs using a human IFN-γ ELISpot plus ALP kit (Mabtech, Stockholm, Sweden). ELISpot plates were washed four times with 200 μL/well Dulbecco’s PBS (Gibco) and were blocked with AIM V media for at least 30 minutes. Two and a half million PBMCs in 100 μL AIM V were stimulated under five conditions including AIM V negative control, SARS-CoV-2 S1 domain (S1) of the spike protein scanning peptide pool (Mabtech), SARS-CoV-2 S2 domain of the spike protein, and the nucleoprotein (S2N) peptide pool (Mabtech), SARS-CoV-2 spike protein, nucleoprotein, membrane protein, open reading frame (ORF)-3a, and ORF-7a proteins (SNMO) peptide pool (Mabtech), and anti-CD3 as a positive control. The final concentration was 2 μg/mL of each peptide. After incubation for 40 hours at 37°C and 5% CO2, cells were removed, and IFN-γ production was determined using biotinylated anti-human IFN-γ mAb 7-B6-1 (1 μg/mL in AIMV; Mabtech) for 2 hours at room temperature, followed by incubation with streptavidin-alkaline phosphatase (1:1,000 in AIM V), and finally treatment with 100 μL ready-to-use BCIP^®^/NBTLiquid substrate (Gibco). After each step, plates were washed five times with distilled water. Emerged spots were counted using an ImmunoSpot analyzer (Cellular Technology Limited, Shaker Heights, OH), and spot quality was checked using ImmunoSpot Software v5.0.9.15. Results were reported as mean and 95% confidence interval (CI) of IFN-γ-producing spot forming units (SFUs) per 10^6^ PBMCs for each peptide pool.

### 2.4 Safety

All patients underwent vital sign measurement and physical examination before vaccination and were then monitored for immediate adverse events (AEs) up to 30 minutes after each vaccination, including local and systemic adverse reactions (**Supplementary Figure 2)**. In addition, solicited AEs were monitored by a phone call on days 3 and 7 after each vaccination (**Supplementary Figure 3)**, and the patients were encouraged to report unsolicited AEs recorded in their diary (**Supplementary Figure 4)**. We then determined the causal association between vaccination and AEs. Participants were also encouraged to contact us to report any possible infections, especially those who developed respiratory symptoms. Those in need of medical attention were asked to visit our facility for further evaluation of adverse reactions or investigation of COVID-19 diagnosis. Nasopharyngeal and oropharyngeal swabs for SARS-CoV-2 RT-PCR were performed if needed to confirm the diagnosis, and treatment was provided according to a standard of care.

### 2.5 Statistical analyses

Continuous variables were expressed as the median with interquartile range (IQR), and categorical variables were presented as absolute and frequencies. The Mann-Whitney U test was performed to compare immunogenicity between KT recipients and controls, and the Wilcoxon signed-rank test was performed to compare within the groups. *P* values <0.05 were considered significant. Statistical analyses were performed with Stata statistical software, version 15 (StataCorp, LLC, College Station, TX). In addition, the distribution of anti-RBD IgG and SARS-CoV-2-specific IFN-γ-producing SFUs/10^6^ PBMCs were expressed as the mean with 95% CI, presented as a dot plot generated with GraphPad Prism 6.0 (GraphPad Software, Inc, San Diego, CA).

### 2.6 Ethics approval

All patients provided written consent. The Institutional Review Board of the Faculty of Medicine, Ramathibodi Hospital, Mahidol University, Bangkok, Thailand, reviewed and approved the study protocol (approval number: MURA2021/242). The study was registered with the Thai Clinical Trials Registry, TCTR20210226002.

## 3. RESULTS

### 3.1 Clinical characteristics of kidney transplant recipients and controls

A prospective study was conducted between April and July 2021. A total of 75 adult patients were vaccinated, including 37 KT recipients and 38 non-transplant controls. Among the former, two were excluded owing to denial participation and prior COVID-19 diagnosis (**Supplementary Figure 1)**. Clinical characteristics of KT recipients are shown in **Table 1**. Among 35 eligible participants, the median age was 50 years (IQR, 42–54), and 60% were male. All (100%) had received a deceased allograft and the majority (97%) had undergone first KT. The median time since transplant was 4.5 years (IQR, 2–9.5). The maintenance immunosuppression regimen included tacrolimus (57%), cyclosporine (29%), corticosteroids (97%), mycophenolic acid (91%) sirolimus (3%), and everolimus (3%). The controls’ median age was 40 years (IQR, 35-44) and 82% were female. There were no immunosuppressive conditions in this group.

**Table 1.** Clinical characteristics of KT recipients (n=35)

| Clinical characteristics, n (%) | N=35 |
| --- | --- |
| Age, years, median (IQR) | 50 (42–54) |
| Male | 21 (60) |
| Asian | 35 (100) |
| Onset from transplant, years |  |
| Within 6 months | 1 (3) |
| 7–12 months | 1 (3) |
| After 1 year | 33 (94) |
| Deceased allograft | 35 (100) |
| First transplant | 34 (97) |
| Immunosuppressants |  |
| Tacrolimus | 20 (57) |
| C<sub>0</sub> level, ng/mL, median (IQR) | 6 (5–7.3) |
| Cyclosporine | 10 (29) |
| C<sub>0</sub> level, ng/mL, median (IQR) | 86 (66–106) |
| Mycophenolate mofetil | 20 (57) |
| Dose, mg/day, median (IQR) | 1,500 (625–2,000) |
| Mycophenolate sodium | 12 (34) |
| Dose, mg/day, median (IQR) | 1080 (720–1080) |
| Sirolimus | 1 (3) |
| Everolimus | 1 (3) |
| Prednisolone | 34 (97) |
| Dose, mg/day, median (IQR) | 5 (5–5) |

IQR, interquartile range; KT, kidney transplant

### 3.2 SARS-CoV-2 specific HMI responses

At 4 weeks after a single dose of the vaccine, the mean (95% CI) titer for anti-RBD IgG was not significantly different compared with before vaccination in all participants (2.4 [95% CI 1.3– 3.4] vs. 1.8 [95% CI 1.3–2.3], p=0.51). Additionally, at 2 weeks post-second dose of the vaccine, a non-significant increase in anti-RBD IgG antibody was observed (7.8 [95% CI 0.2–15.5] vs. 1.8 [95% CI 1.3–2.3], p=0.07; **Figure 1)**. In comparison with non-transplant controls, the mean (95% CI) anti-RBD IgG titer was significantly lower in the KT group at 2 weeks post-second dose (2691 [95% CI 1581–3802] vs. 7.8 [95% CI 0.2–15.5], p<0.001; **Table 2)**. The mean (95% CI) percentages of surrogate virus neutralization antibody inhibition was also significantly lower in the KT group at 2 weeks post-second dose (2 [95% CI −1–6] vs. 71 [95%CI 61–81], p<0.001); **Figure 2**).

**Table 2.**
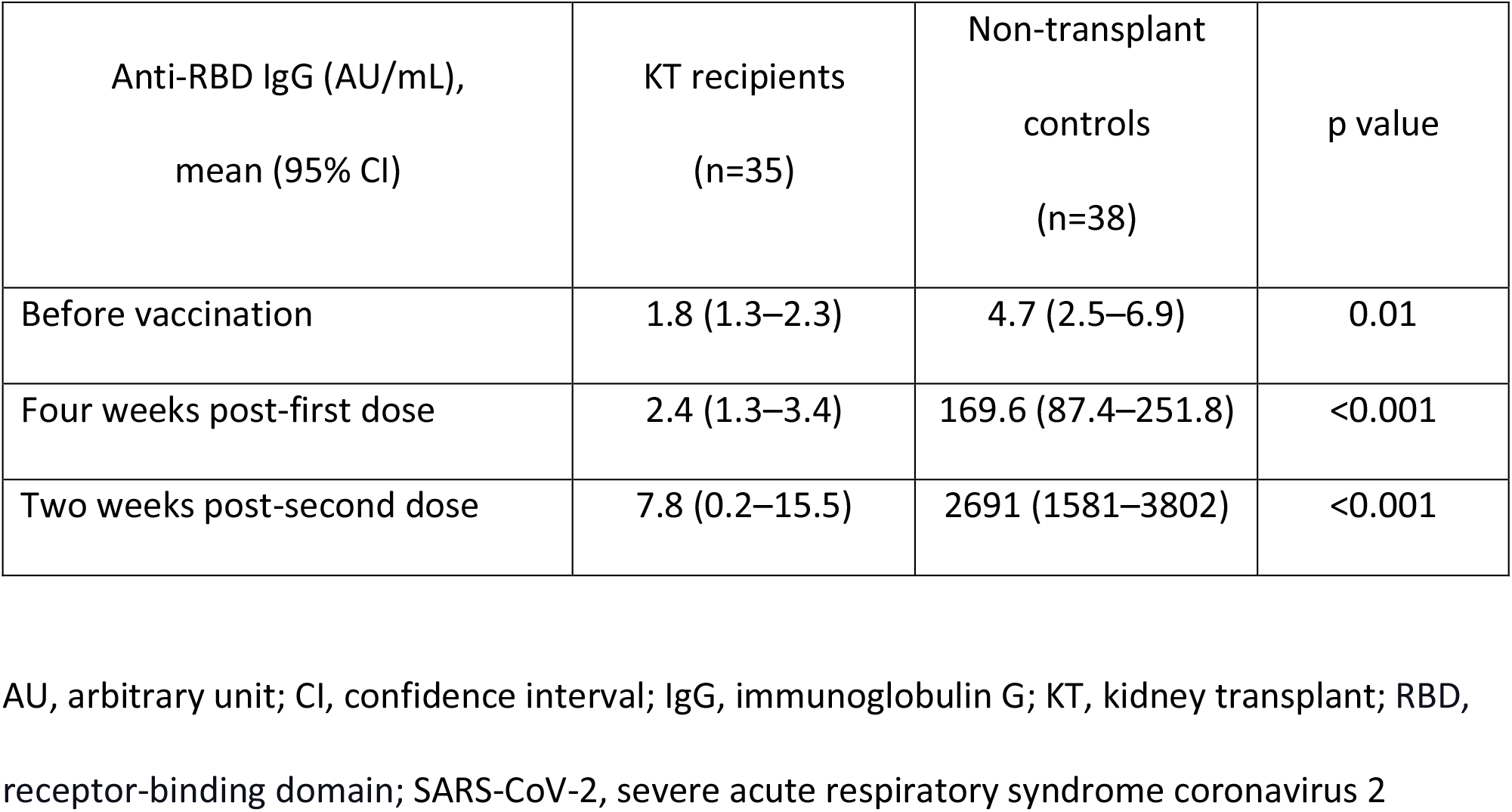
SARS-CoV-2-specific HMI responses represented by anti-RBD IgG in KT recipients and non-transplant controls vaccinated with inactivated SARS-CoV-2 vaccine

**Figure 1.**
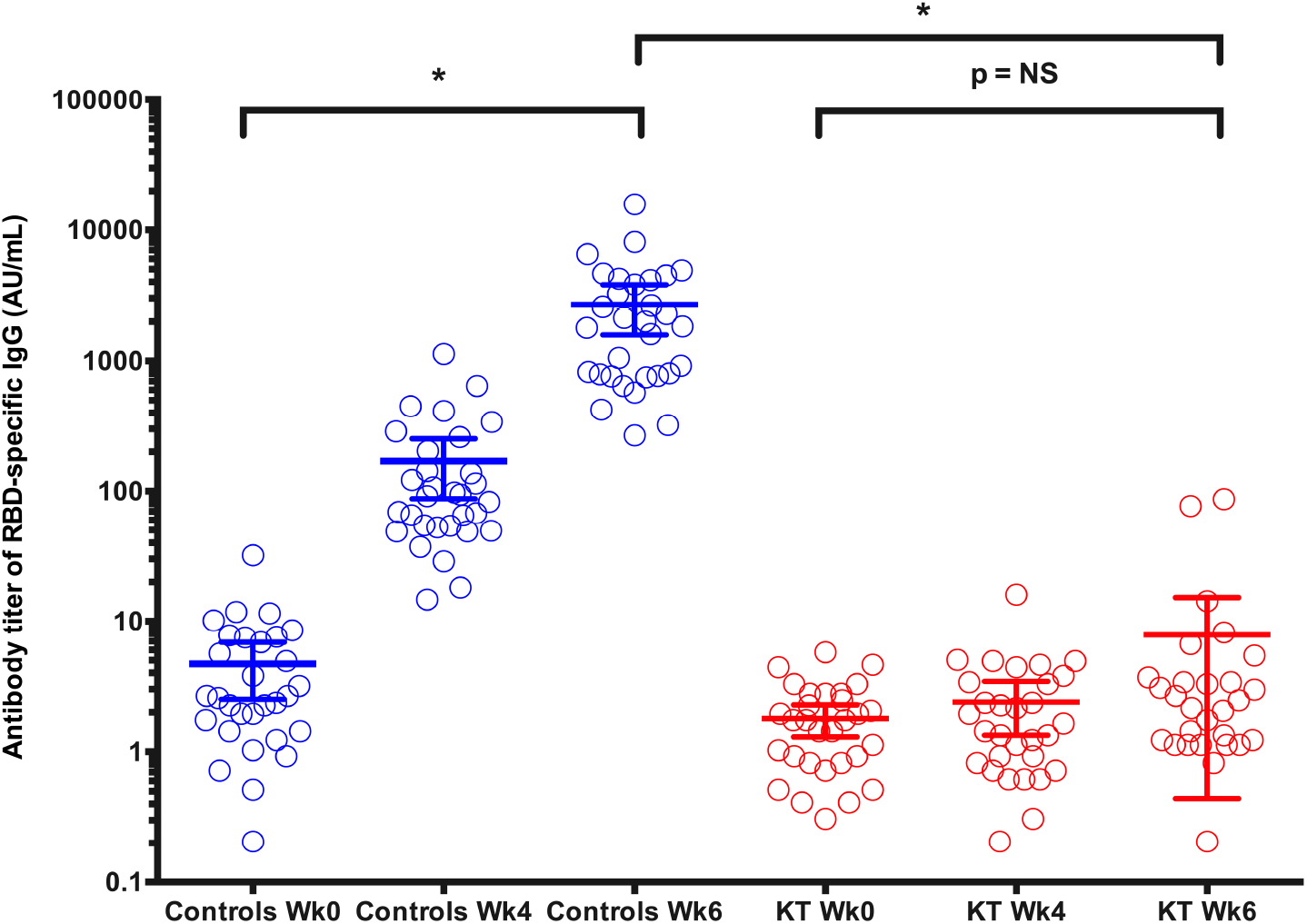
The prevalence of SARS-CoV-2 RBD-specific IgG antibody titer before, 4 weeks post-first dose and 2 weeks post-second dose in non-transplant controls and KT recipients. * P value < 0.05.

**Figure 2.**
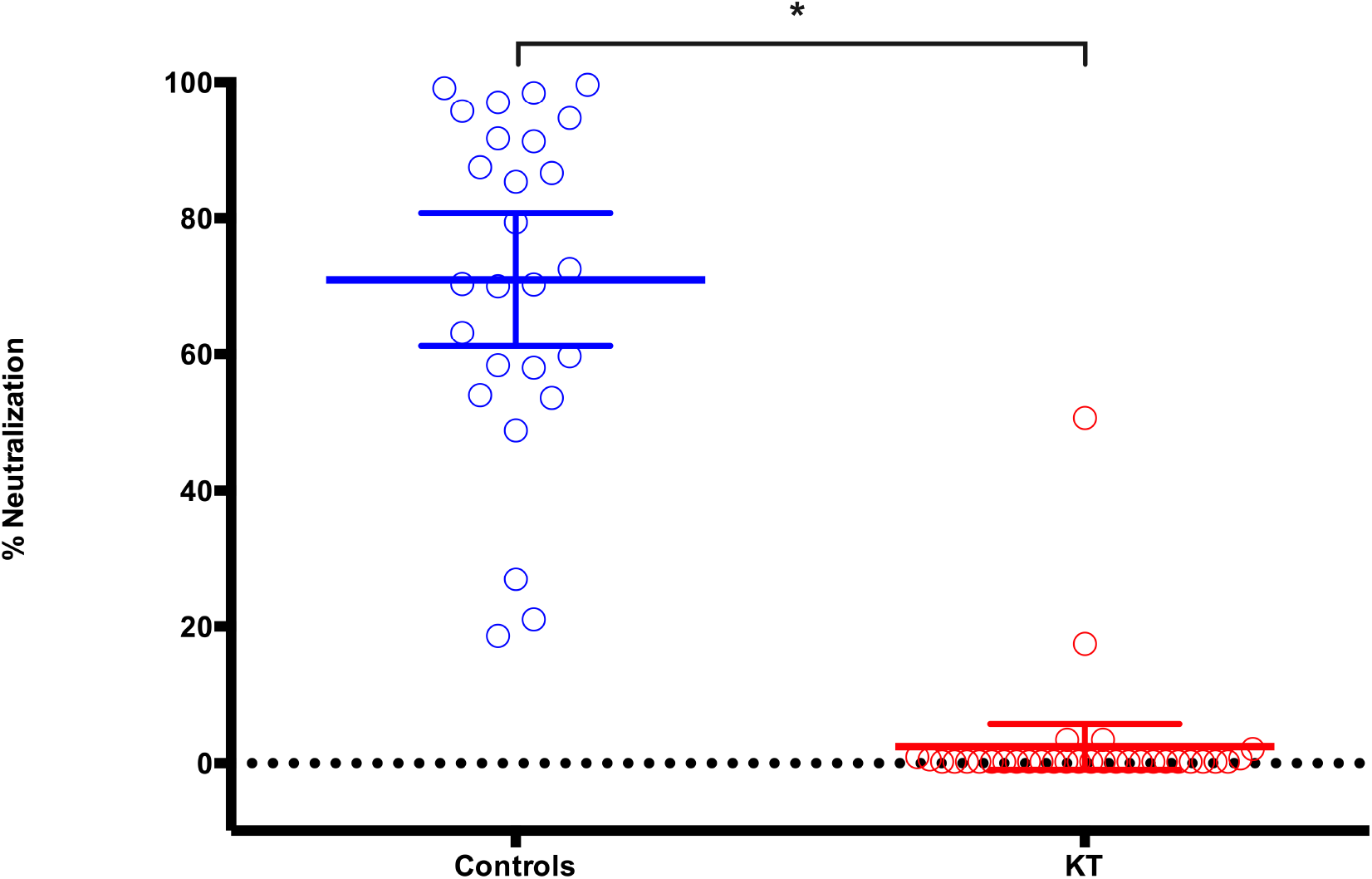
The prevalence of surrogate virus neutralization antibody inhibition at 2 weeks post-second dose in non-transplant controls and kidney transplant recipients. * P value < 0.05.

### 3.3 SARS-CoV-2 specific CMI responses

At 4 weeks after a single dose of the vaccine, mean (95% CI) S1 and SNMO-specific T-cell responses were not significantly different compared with before vaccination in all participants **(Figure 3)**. However, S2N-specific T-cell responses were significantly decreased compared with before vaccination (10 [95% CI 3–17] vs. 20 [95% CI −4–44] specific T-cells/10^6^ PBMCs, p=0.008).

**Figure 3.**
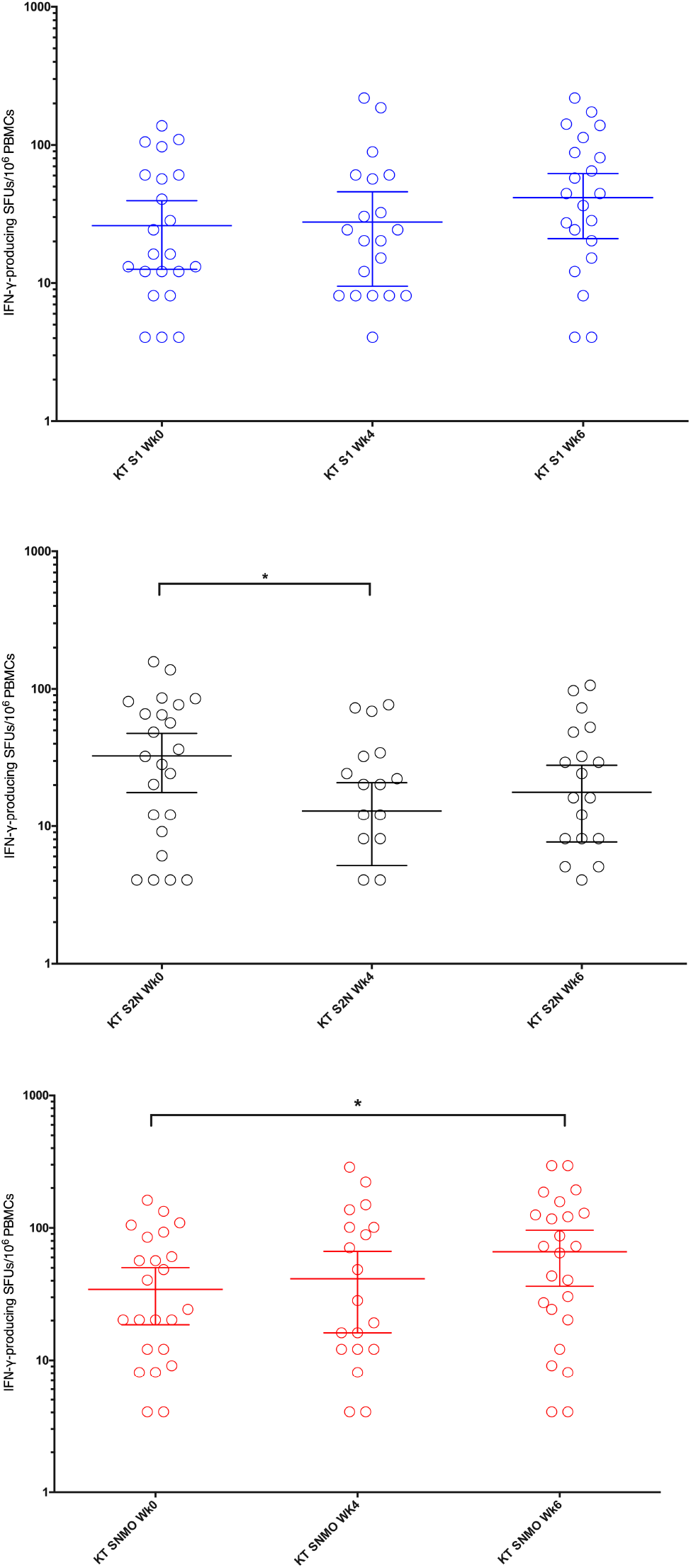
SARS-CoV-2-specific IFN-γ-producing T-cell responses reactive to the S1 protein, S2N protein, and the SMNO protein responses detected by IFN-γ ELISpot assay before vaccination, 4 weeks post-first dose and 2 weeks post-second dose in KT recipients. * P value < 0.05. **Abbreviations:** IFN-γ, interferon-γ; SFU, spot forming unit; PBMC, peripheral blood mononuclear cell; S, spike glycoprotein; S1, S1 domain of spike protein; S2N, spike and nucleoproteins; SNMO: peptide pool of spike protein, nucleoprotein, membrane protein and open reading frame proteins

At 2 weeks post-second dose of the vaccine, mean (95% CI) SNMO-specific T-cell responses were significantly increased compared with before vaccination (66 [95% CI 36–99] vs. 34 [95% CI 19–50] specific T-cells/10^6^ PBMCs, p=0.02). However, S1 and S2N-specific T-cell responses were not significantly different compared with baseline (p=NS; **Table 3)**.

**Table 3.**
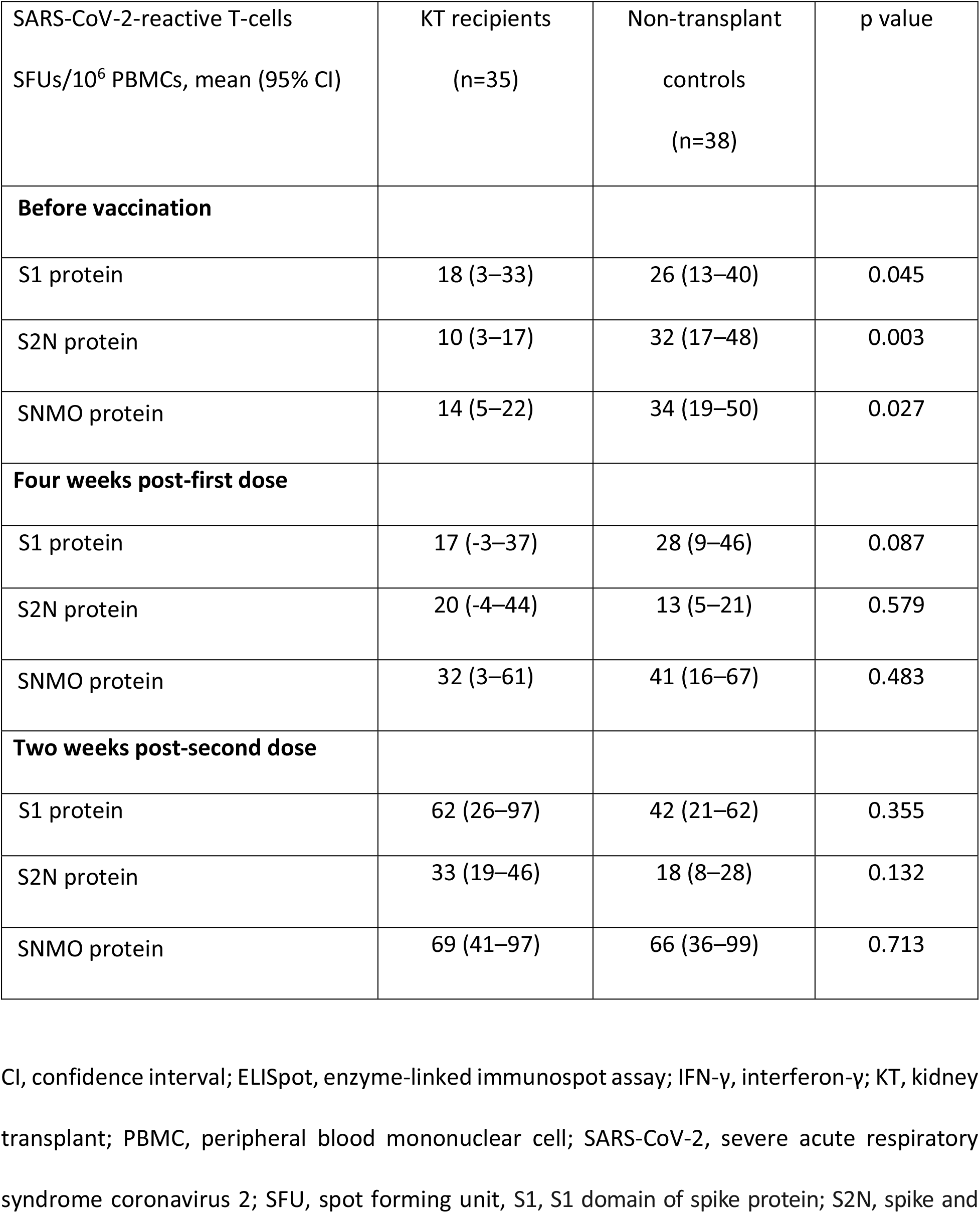
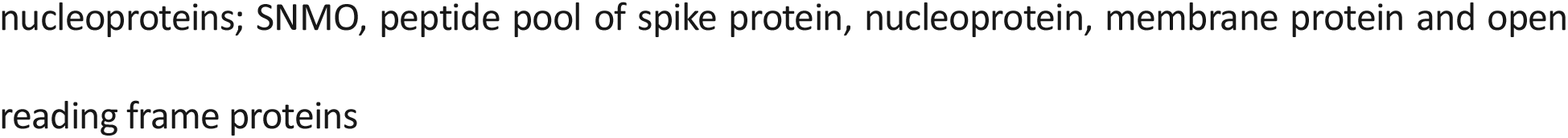
SARS-CoV-2-specific T-cell responses assessed by the IFN-γ ELISpot assay in KT recipients and non-transplant controls vaccinated with inactivated SARS-CoV-2 vaccine

Two representative KT recipients without and with SARS-CoV-2-specific IFN-γ-producing T-cell responses to the S1 protein, S2N protein, and the SMNO protein detected by IFN-γ ELISpot assay at 2 weeks post-second dose of inactivate COVID-19 vaccine; **Figure 4**.

**Figure 4.**
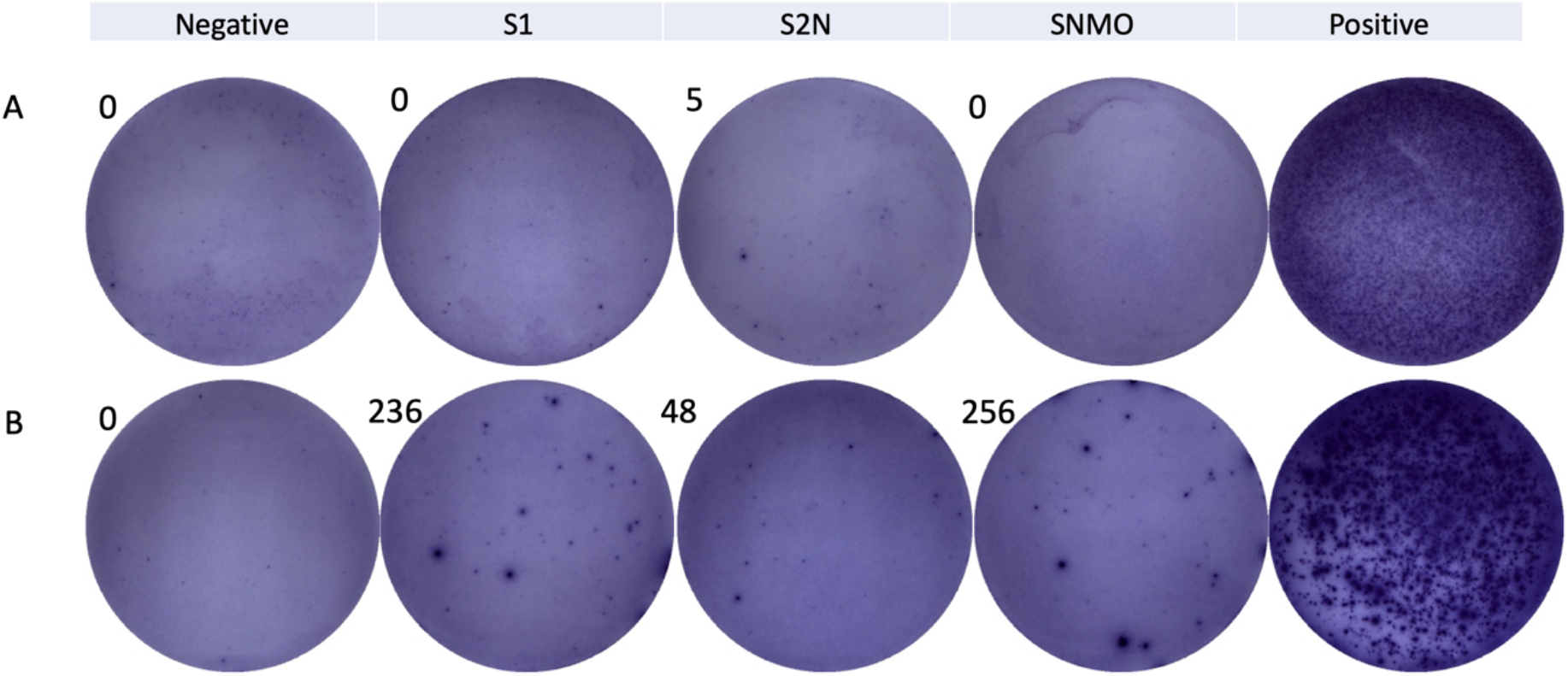
Two representative KT recipients without (A) and with (B) SARS-CoV-2-specific IFN-γ-producing T-cell responses to the S1 protein, S2N protein, and the SMNO protein detected by IFN-γ ELISpot assay at 2 weeks post-second dose of inactivate COVID-19 vaccine. The number in the upper left corner of each well indicates the number of spot-forming units in each well. Positive and negative results were defined according to the manufacturer’s recommendations. **Abbreviations:** IFN-γ, interferon-γ; S, spike glycoprotein; S1, S1 domain of spike protein; S2N, spike and nucleoproteins; SNMO: peptide pool of spike protein, nucleoprotein, membrane protein and open reading frame proteins

### 3.4 Safety

No severe local or systemic AEs were observed immediately within 30 minutes after each dose of the vaccine. Solicited AEs are presented in **Supplementary Table 1**. On day 3 after the first dose, 16 (46%) of participants reported no AEs, and the remainder reported AEs including fever (17%), pain at the injection site (14%), sleepiness (9%), muscle aches (6%), increased appetite (3%) and others (6%). Two (6%) unsolicited AEs were reported, including asthmatic attack (n=1) and subconjunctival hemorrhage (n=1). Both were evaluated and deemed not related to the vaccine. On day 7 after the first dose, the majority of participants (94%) reported no AEs. On day 3 after the second dose, 18 (51%) reported no AEs. However, the remainder reported the following: fever (11%), pain at the injection site (9%), muscle aches (9%), sleepiness (14%), and others (6%). On day 7 after the second dose, 34 (97%) reported no AEs. No acute rejection episodes occurred in those who were fully vaccinated. All observed AEs were mild (grade 1) in severity and recovery occurred within 48–72 hours, except for one patient with asthma who required an outpatient visit and was prescribed a bronchodilator inhaler (grade 3).

## 4. DISCUSSION

We herein present a pilot study investigating immunogenicity, focusing on immune responses specific to SARS-CoV-2 after vaccination with an inactivated whole SARS-CoV-2 vaccine administered at an interval of 28 days in patients who previously underwent KT and were receiving immunosuppressive agents. The development of HMI, indicated by anti-RBD IgG titers and the percentage of neutralizing antibodies inhibition, was not adequately achieved after two doses of vaccine and was poor compared with non-transplant individuals. Conversely, a significant increasing trend was observed for CMI, quantified by SARS-CoV-2-specific IFN-γ-producing T-cells after stimulation with SARS-CoV-2 mixed peptides. In addition, the short-term safety and clinical profile was acceptable but warrant further study.

SOT recipients are considered to have comorbidities and are at greater risk of severe respiratory tract disease.^13^ Among several COVID-19 vaccines available, a two-dose regimen of SARS-CoV-2 mRNA vaccine revealed immunogenicity in SOT recipients at 40%, and a third dose was required to boost a more significant response to 68%.^8–10, 14^ KT recipients receiving adenovirus-vectored vaccine could still be vulnerable to infection, reflecting a possible inadequate immune response in a small recent study.^15^ Our study also confirmed a weak HMI response and a optimal CMI production. Although no threshold has been established for protective immunity, antibody levels and the amount of neutralizing property of those were well below those observed in the vaccinated immunocompetent group. We also observed that the mean RBD-specific IgG antibody titer and neutralizing antibody inhibition in our participants was lower than that of immunocompetent patients vaccinated with CoronaVac^®^ in phase 1 and 2 studies.^16^ However, neutralizing antibody measured by plaque reduction test is believed to be a valid test to assess protective immunity, although an appropriate cut-off value to determine those with sufficient neutralizing titer has not yet been established and requires a post-marketing study to prove its effectiveness.^17^

We are among the first to report the induction of CMI, focusing on quantification of T-cells stimulated with SARS-CoV-2 peptides in a post-transplant immunocompromised population. Although SARS-CoV-2-reactive T-cell responses to an isolated S1 domain of the spike protein alone or combined with nucleocapsid protein were not significantly increased after complete vaccination, we observed increased responses to mixed peptides (SNMO) after the second dose.

The development of responses to SARS-CoV-2 mixed peptides could be due to a natural characteristic of the whole virus we selected. S1-specific T-cell responses would be more significant among those receiving mRNA-based or viral vector vaccines which targeted the S1 domain of the spike protein. Moderate generation of IFN-γ-producing T-cell responses among those receiving 3–6 μg inactivated virus-containing vaccines were 3.4 and 1.2 SFU/10^6^ PBMCs, respectively (the former produced more) in a relatively new cohort, which was lower compared with our results even in those with intact immunity.^16^ Our study also revealed a relatively comparable CMI after immunization to the control group and supported that 3 μg inactivated virus-containing vaccine to robust CMI should be adequate. The wash step in the ELISpot assay could subside an immunosuppressive effect to the T cells and create an intact response. CMI response was also detectable after mRNA-based vaccination in SOT recipients in a recent study.^18^ We believe an intact CMI induced by memory T-cells is essential and could be activated during natural infection, thus decreasing the severity of the disease.

Several factors could diminish immune responses after vaccination in SOT recipients, not least the fact that renal allografts must be maintained by immunosuppressive agents. Mycophenolate mofetil treatment of greater than 1 g per day, as in our recipients, has been reported in the literature as a critical factor to blunt an immune response. The virus contained in the CoronaVac^®^ vaccine should be more than 3 mg to produce adequate immunogenicity in patients receiving immunosuppressants. These data are compatible with immunogenicity generated following a standard dose of inactivated influenza vaccine in KT recipients, which revealed lower antibody titers than non-transplant immunocompromised populations such as patients living with HIV or end-stage renal disease.^19^ Therefore, influenza vaccine formulations with a higher dose of hemagglutinin is encouraged for those in need, such as elderly individuals or SOT recipients, to generate a stronger immune response compared with the standard dose.^20–22^ A recent study evaluating immunogenicity after triple doses of an mRNA COVID-19 vaccine in SOT recipients indicated that this approach could achieve an optimal response and be promising.^14^ There is the possibility that additional vaccine doses would be needed, or switching to another vaccine platform could be intriguing. *Heterogenous vaccine* studies have been more focused on investigation of the immunocompetent population while our specific post-transplant population is often excluded from the study. Although immunosuppressive agents could blunt our patients’ immunity, we observed more HMI effects than CMI. The responses of CMI in KT recipients were not statistically significant compared to the controls could be explained by a wash step of ELISpot assay, which attempts to decrease the effect of T cell immunosuppressants on their responses.

Safety is another issue of concern among SOT recipients. Adverse reactions during the early period were reported to be mild, confirmed by a large cohort prospective study of mRNA vaccine provided to SOT recipients. The most common AE reported in a phase 1/2 study of the inactivated whole virus vaccine was injection-site pain, reported by approximately one in five participants; this was higher than the rate reported in our study of 14%.^16^ Our study confirmed that only minimal and mild adverse reactions were observed following vaccination in these unexplored populations. However, immediate and short-term AEs are tolerable. Long-term adverse events and allograft profiles such as allograft rejection require further follow-up.

Limitations of this study include the small sample size. Future large-scale studies are needed to confirm our findings and further explore predictors of inadequate immune responses in these specific populations. The strength of this study is it represents one of the first studies to investigate immunogenicity and safety in SOT recipients vaccinated with an inactivated SARS-CoV-2 vaccine. Although poor anti-RBD antibody and surrogate neutralization antibody responses were observed compared with immunocompetent individuals, the assumption of inadequate humoral responses cannot yet be completely elucidated, as further studies using standardized plaque reduction neutralization tests are necessary to define a better cut-off antibody titer that correlates well with neutralization. However, we instead attempted to assess CMI, which is believed to boost a prolonged protective memory response in our susceptible patients. However, the most important thing is adherence to strict basic infection prevention measures remains crucial after immunization.

So far, research focused on the effectiveness of COVID-19 vaccines in SOT recipients has not been fully explored. Our study could not report the effectiveness of this vaccine in preventing natural infection because of the short follow-up period after vaccination. Furthermore, vaccine effectiveness varies depending on the study population, the dynamics of local virus transmission, the dominance of variants of concern, and healthcare resources. Thus, post-marketing investigations will be required to determine the efficacy of vaccination in SOT recipients. Allograft safety profiles and long-term data on safety also need to be followed up. Nevertheless, we believe our findings could provide preliminary data on SARS-CoV-2 immune responses following whole virus SARS-CoV-2 vaccination and be beneficial in designing an appropriate strategy for vaccination in SOT recipients.

Our study revealed that KT recipients develop weak antibody responses and their neutralizing effect to the spike protein, but with marginal SARS-CoV-2-specific T-cell responses after completing a two-dose course of inactivated SARS-CoV-2 vaccine with acceptable adverse reactions and favorable short-term outcomes. However, this population could remain unprotected and vulnerable to severe consequences of SARS-CoV-2 infection. Therefore, consideration of providing a booster dose with the same platform or switching to another heterogeneous vaccine should be verified in future studies. An optimal vaccine strategy could potentially improve immune responses in SOT recipients and those patients receiving immunosuppressants.

## Data Availability

Data will be made available upon appropriate request.

## Acknowledgments

This study received a grant from the National Research Council of Thailand (NRCT), Ministry of Higher Education, Science, Research, and Innovation of Thailand in collaboration with the Department of Medical Services, Ministry of Public Health of Thailand (102912). The authors would like to thank the Ramathibodi Transplant Infectious Diseases (RTID) Study Group. The group members are Vasant Sumethkul, Somnuek Domrongkitchaiporn, Bunyong Phakdeekitcharoen, Chagriya Kitiyakara, Sinee Disthabanchong, Atiporn Ingsathit, Punlop Wiwattanathum, and Ithikorn Spanuchart. We offer our gratitude to Pongsan Chatsangcharoen and Parnwas Pinnobphun for their laboratory support. We are also grateful for infectious diseases nurses and supportive staffs at the Division of Infectious Diseases, Department of Medicine, and Clinical Research Center, Faculty of Medicine Ramathibodi Hospital, Mahidol University, Bangkok, Thailand.

## Supplementary figures

**Supplementary Figure 1.**
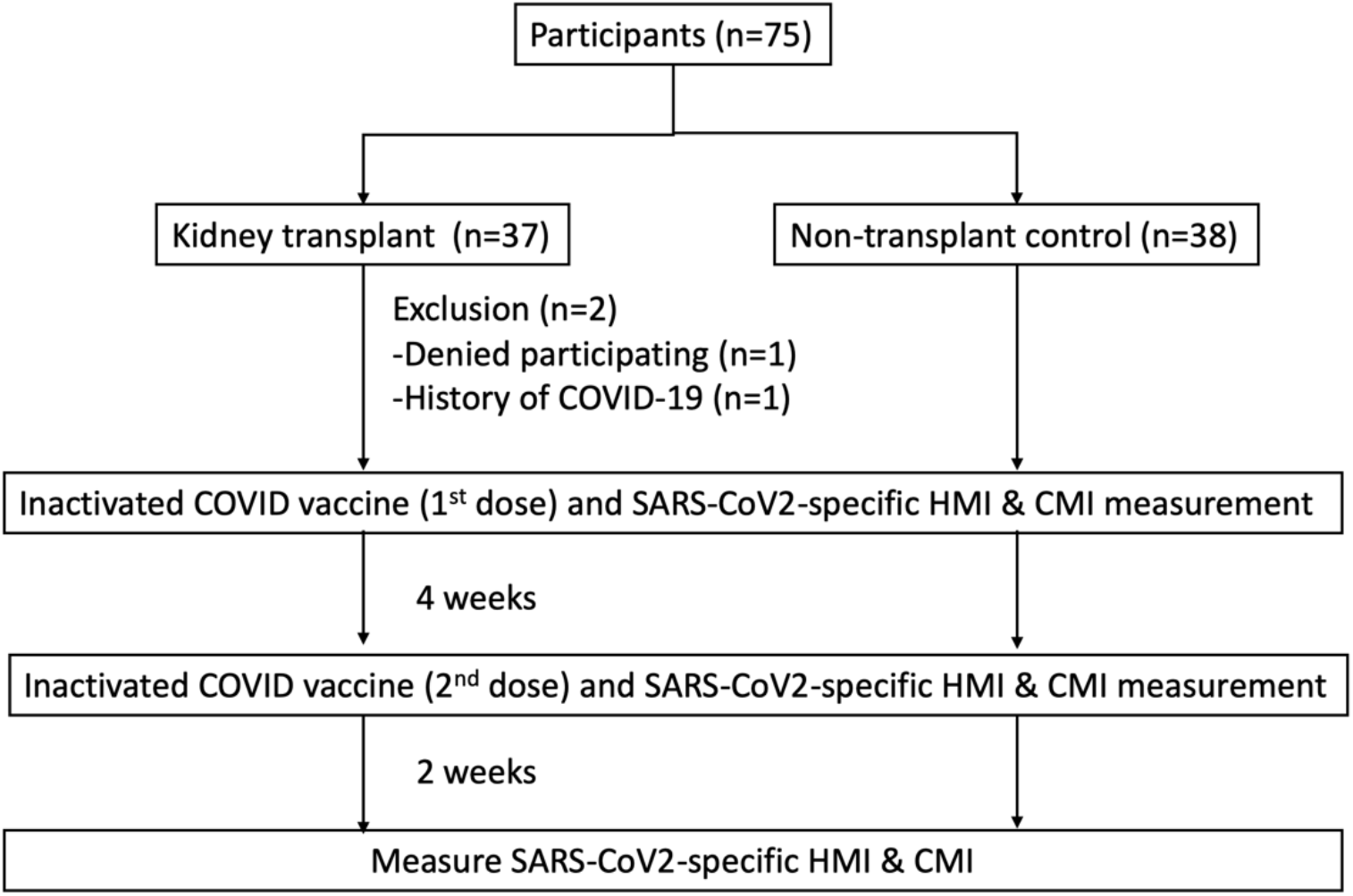
Study design

**Supplementary Figure 2.**
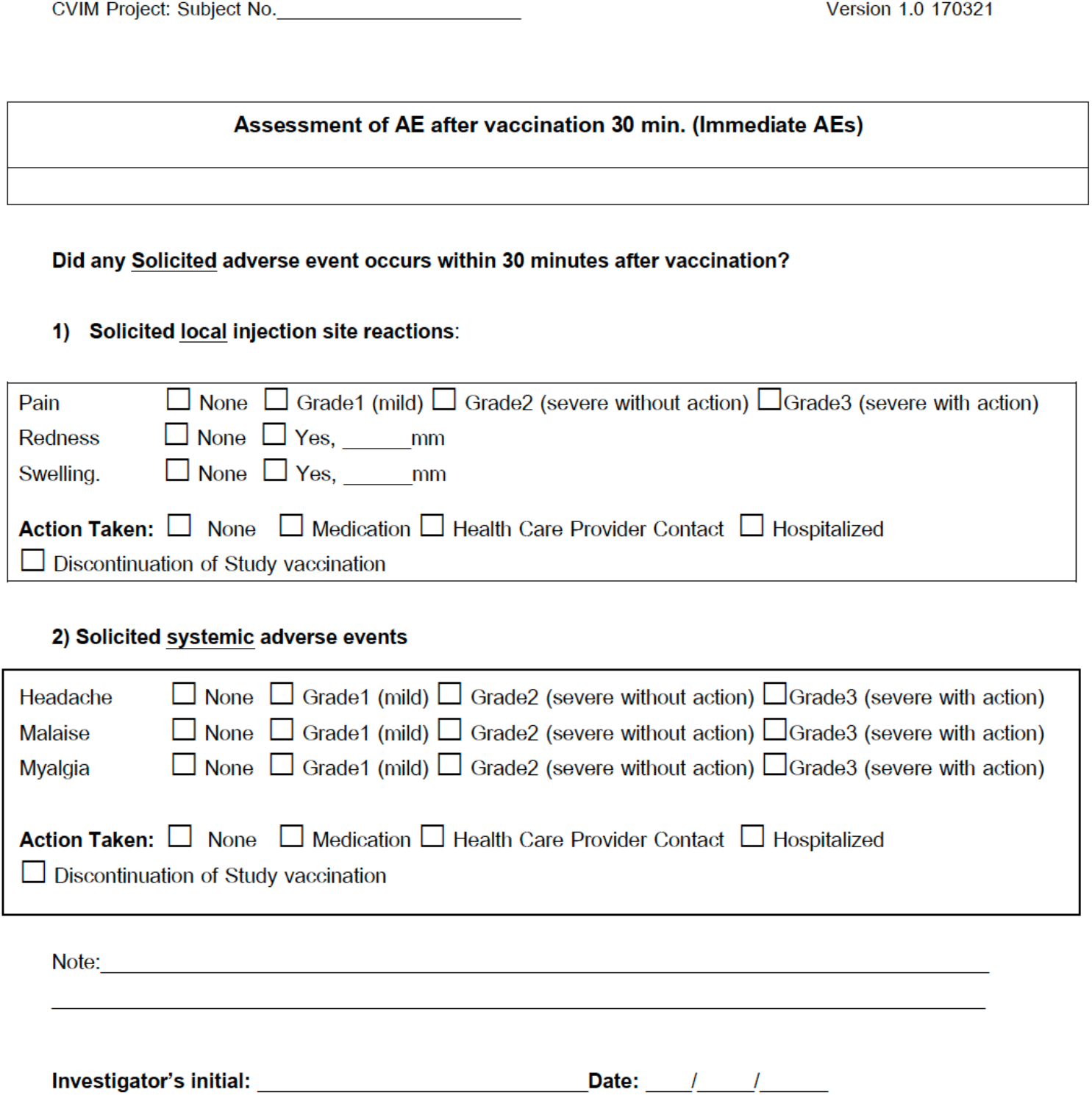
Assessment sheet of immediate adverse events after vaccination

**Supplementary Figure 3.**
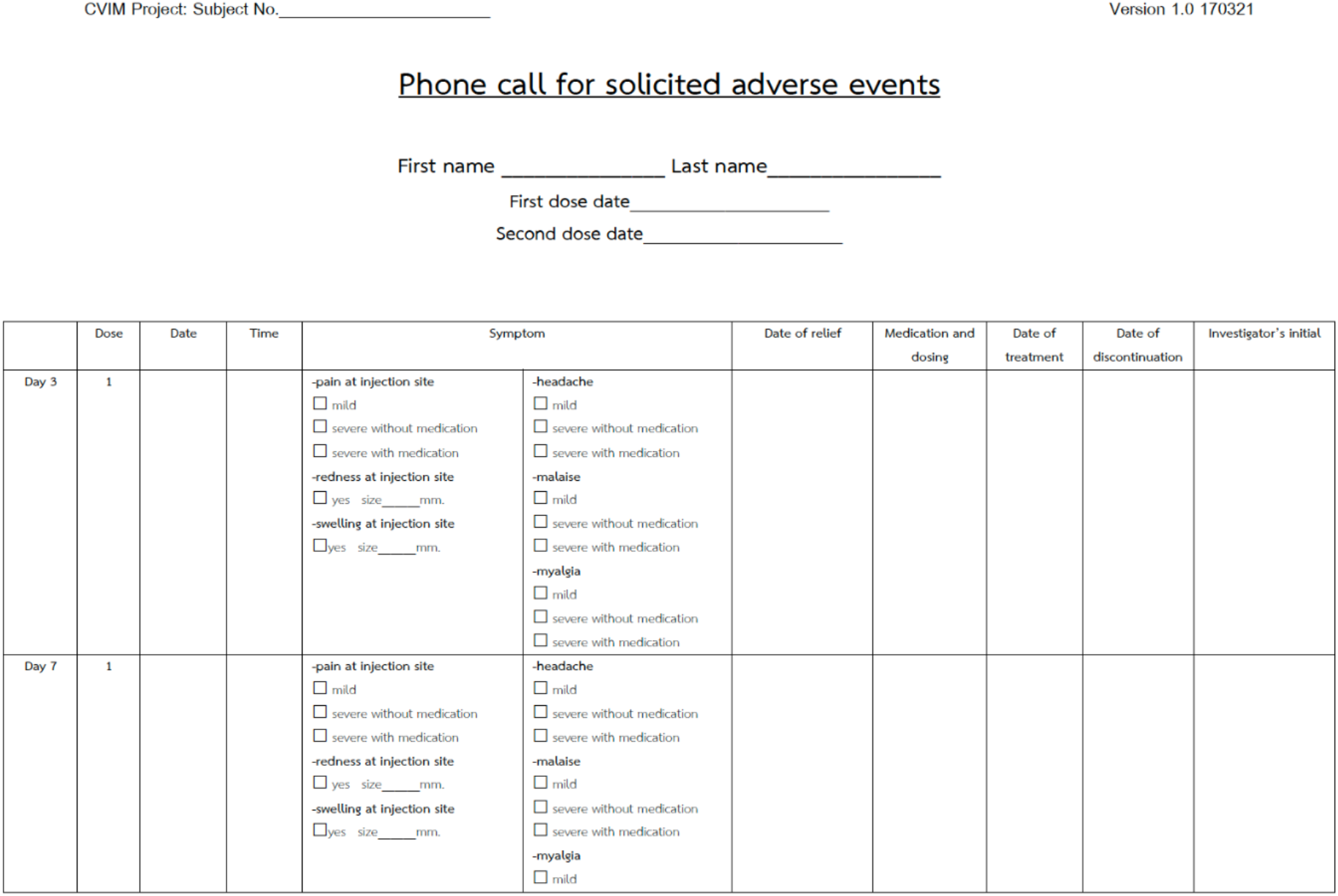
Assessment sheet of solicited adverse events after vaccination

**Supplementary Figure 4.**
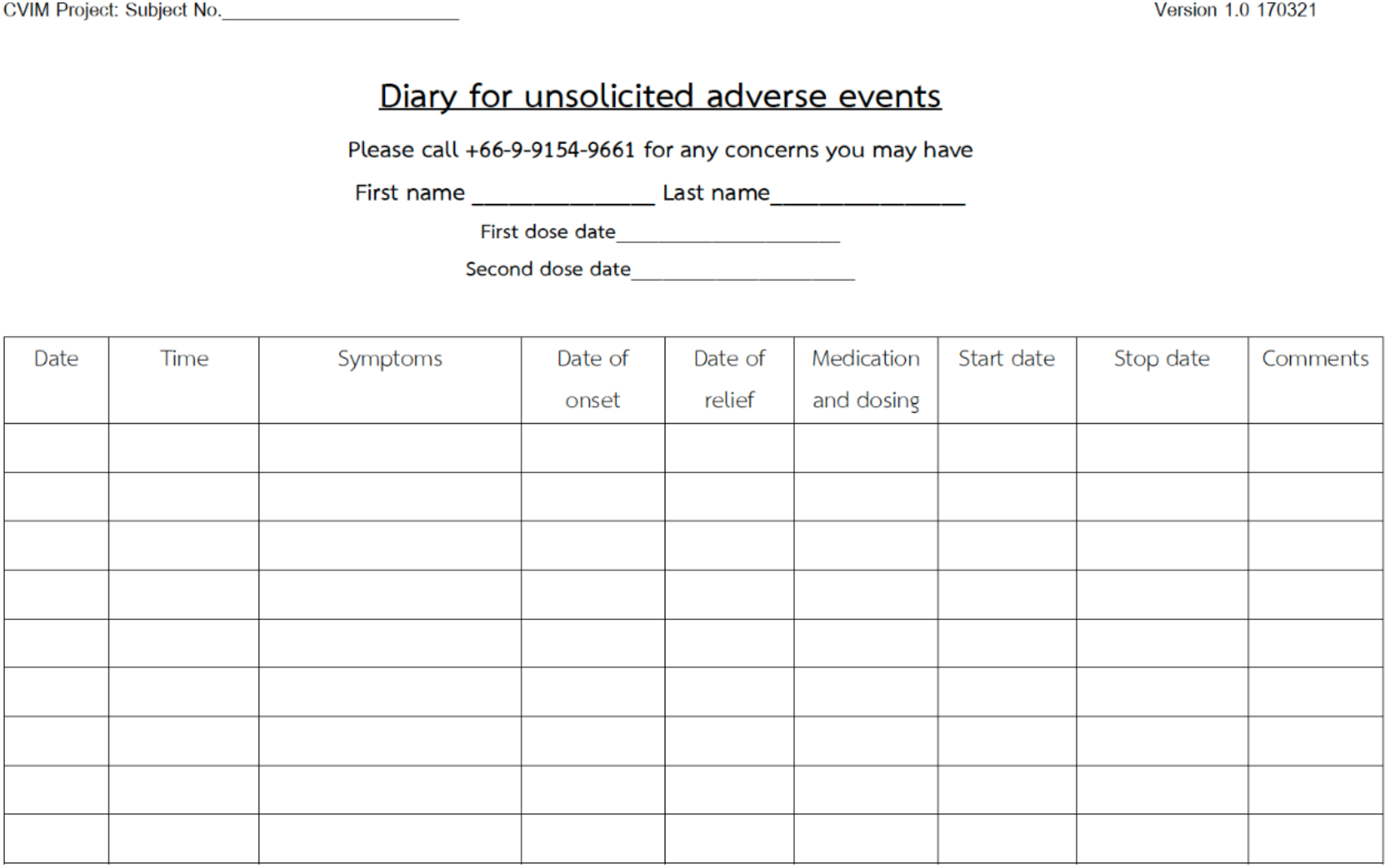
Assessment sheet of unsolicited adverse events after vaccination

**Supplementary Table 1.** Solicited adverse events at days 3 and 7 after vaccination

| N (%) | Days post-first dose |  | Days post-second dose |  |
| --- | --- | --- | --- | --- |
|  | Day 3 | Day 7 | Day 3 | Day 7 |
| No adverse events | 16 (46) | 33 (94) | 18 (51) | 34 (97) |
| Pain at the injection site | 5 (14) | 0 | 3 (9) | 0 |
| Muscle aches | 2 (6) | 0 | 3 (9) | 0 |
| Increased appetite | 1 (3) | 0 | 0 | 0 |
| Fever | 6 (17) | 0 | 4 (11) | 0 |
| Sleepiness | 3 (9) | 0 | 5 (14) | 1 (3) |
| Others | 2 (6) | 2 (6) | 2 (6) | 0 |

## Abbreviations

ACE2: angiotensin-converting enzyme 2
AE: adverse events
CI: confidence interval
CMI: SARS-CoV-2-specific cell-mediated immunity
COVID-19: coronavirus disease 2019
ELISpot: enzyme-linked immunospot assay
HMI: SARS-CoV-2-specific humoral immunity
IFN-γ: interferon-γ
IgG: immunoglobulin G
IQR: interquartile range
KT: kidney transplant
M: SARS-CoV-2 membrane protein
mRNA: messenger ribonucleic acid
N: SARS-CoV-2 nucleoprotein
ORF: open reading frame
PBMC: peripheral blood mononuclear cell
RBD: receptor binding domain
RT-PCR: reverse-transcription polymerase chain reaction
S: SARS-CoV-2 spike glycoprotein
SARS-CoV-2: severe acute respiratory syndrome coronavirus 2 SFU spot forming units
SMNO: SARS-CoV-2 spike protein, nucleoprotein, membrane protein and ORF-3a, and ORF-7a proteins
SOT: solid organ transplantation
sVNT: SARS-CoV-2 surrogate virus neutralization test

